# Phenotype of SARS-CoV-2-specific T-cells in COVID-19 patients with acute respiratory distress syndrome

**DOI:** 10.1101/2020.04.11.20062349

**Authors:** Daniela Weiskopf, Katharina S. Schmitz, Matthijs P. Raadsen, Alba Grifoni, Nisreen M.A. Okba, Henrik Endeman, Johannes P.C. van den Akker, Richard Molenkamp, Marion P.G. Koopmans, Eric C.M. van Gorp, Bart L. Haagmans, Rik L. de Swart, Alessandro Sette, Rory D. de Vries

## Abstract

SARS-CoV-2 has been identified as the causative agent of a global outbreak of respiratory tract disease (COVID-19). In some patients the infection results in moderate to severe acute respiratory distress syndrome (ARDS), requiring invasive mechanical ventilation. High serum levels of IL-6 and an immune hyperresponsiveness referred to as a ‘cytokine storm’ have been associated with poor clinical outcome. Despite the large numbers of cases and deaths, information on the phenotype of SARS-CoV-2-specific T-cells is scarce. Here, we detected SARS-CoV-2-specific CD4^+^ and CD8^+^ T cells in 100% and 80% of COVID-19 patients, respectively. We also detected low levels of SARS-CoV-2-reactive T-cells in 20% of the healthy controls, not previously exposed to SARS-CoV-2 and indicative of cross-reactivity due to infection with ‘common cold’ coronaviruses. Strongest T-cell responses were directed to the surface glycoprotein (spike, S), and SARS-CoV-2-specific T-cells predominantly produced effector and Th1 cytokines, although Th2 and Th17 cytokines were also detected. Collectively, these data stimulate further studies into the role of T-cells in COVID-19, support vaccine design and facilitate the evaluation of vaccine candidate immunogenicity.

**Summary:** COVID-19 is associated with lymphopenia and ‘cytokine storm’, but there is a scarcity of information on specific cellular immune responses to SARS-CoV-2. Here, we characterized SARS-CoV-2-specific CD4^+^ and CD8^+^ T-cells in patients hospitalized with acute respiratory distress syndrome (ARDS).

## Introduction

A novel coronavirus, named SARS-CoV-2, has been identified as the causative agent of a global outbreak of respiratory tract disease, referred to as COVID-19 (Chan et al., 2020; Huang et al., 2020). On May 1^st^, over 3 million cases and more than 230,000 deaths were reported globally. COVID-19 is characterized by fever, cough, dyspnea and myalgia (Huang et al., 2020), but in some patients the infection results in moderate to severe acute respiratory distress syndrome (ARDS), requiring invasive mechanical ventilation for a period of several weeks. COVID-19 patients may present with lymphopenia (Chen et al., 2020; Huang et al., 2020), but the disease has also been associated with immune hyperresponsiveness referred to as a ‘cytokine storm’ (Mehta et al., 2020). A transient increase in co-expression of CD38 and HLA-DR by T-cells, a phenotype of CD8^+^ T-cell activation in response to viral infection, was observed concomitantly (Xu et al., 2020). This increase in both CD4^+^ and CD8^+^ CD38^+^HLA-DR^+^ T-cells preceded resolution of clinical symptoms in a non-severe, recovered, COVID-19 patient (Thevarajan et al., 2020).

Despite the large numbers of cases and deaths, there is a scarcity of information on the phenotype of SARS-CoV-2-specific T-cells. Spike surface glycoprotein (S-), M-and NP-specific T-cells were detected in PBMC from convalescent COVID-19 patients (Ni et al., 2020). Virus-specific T-cells have also been detected after exposure to the related SARS-CoV and MERS-CoV, although few studies have characterized cellular responses in human patients. For SARS-CoV-specific CD4^+^ T-cells it was reported that the S glycoprotein accounted for nearly two thirds of T-cell reactivity, N and M also accounted for limited reactivity (Li et al., 2008). For MERS-CoV-specific CD4^+^ T-cells responses targeting S, N and a pool of M and E peptides have been reported (Zhao et al., 2017).

Here, we stimulated peripheral blood mononuclear cells (PBMC) from ten COVID-19 patients with ARDS, collected up to three weeks after admission to the intensive care unit (ICU), with MegaPools (MP) of overlapping or prediction-based peptides covering the SARS-CoV-2 proteome (Grifoni et al., 2020). We detected SARS-CoV-2-specific CD4^+^ and CD8^+^ T-cells in 10/10 and 8/10 COVID-19 patients, respectively. Peptide stimulation of healthy control (HC) PBMC samples collected before the outbreak in most cases resulted in undetectable responses, although some potential cross-reactivity due to infection with ‘common cold’ coronaviruses was observed. SARS-CoV-2-specific T-cells predominantly produced effector and Th1 cytokines, although Th2 and Th17 cytokines were also detected.

## Results & Discussion

We included ten COVID-19 patients with moderate to severe ARDS in this study. All patients were included in the study shortly after ICU admission; self-reported illness varied between 5 and 14 days before inclusion (Figure 1a). Patients were between 49 and 72 years old (average 58.9 ± 7.2 years) and of mixed gender (4 female, 6 male). All patients tested SARS-CoV-2 positive by RT-PCR and were ventilated during their stay at the ICU. At the time of writing, 5 patients were transferred out of the ICU (case 1, 2, 4, 6 and 7), 3 patients were still in follow-up (case 5, 9 and 10), 1 patient was discharged (case 8) and 1 patient was deceased (case 3). Case 4 was deceased 4 days after transfer out of the ICU. Patients were treated with lung protective ventilation using the higher PEEP/lower FiO_2_ table of the ARDSnet and restrictive volume resuscitation. They received antibiotics as a part of a treatment regimen aimed at selective decontamination of the digestive tract. Furthermore, all patients received chloroquine, lopinavir, ritonavir and / or corticosteroids for a brief period of time around admission to the ICU (Figure 1a).

**Figure 1.**
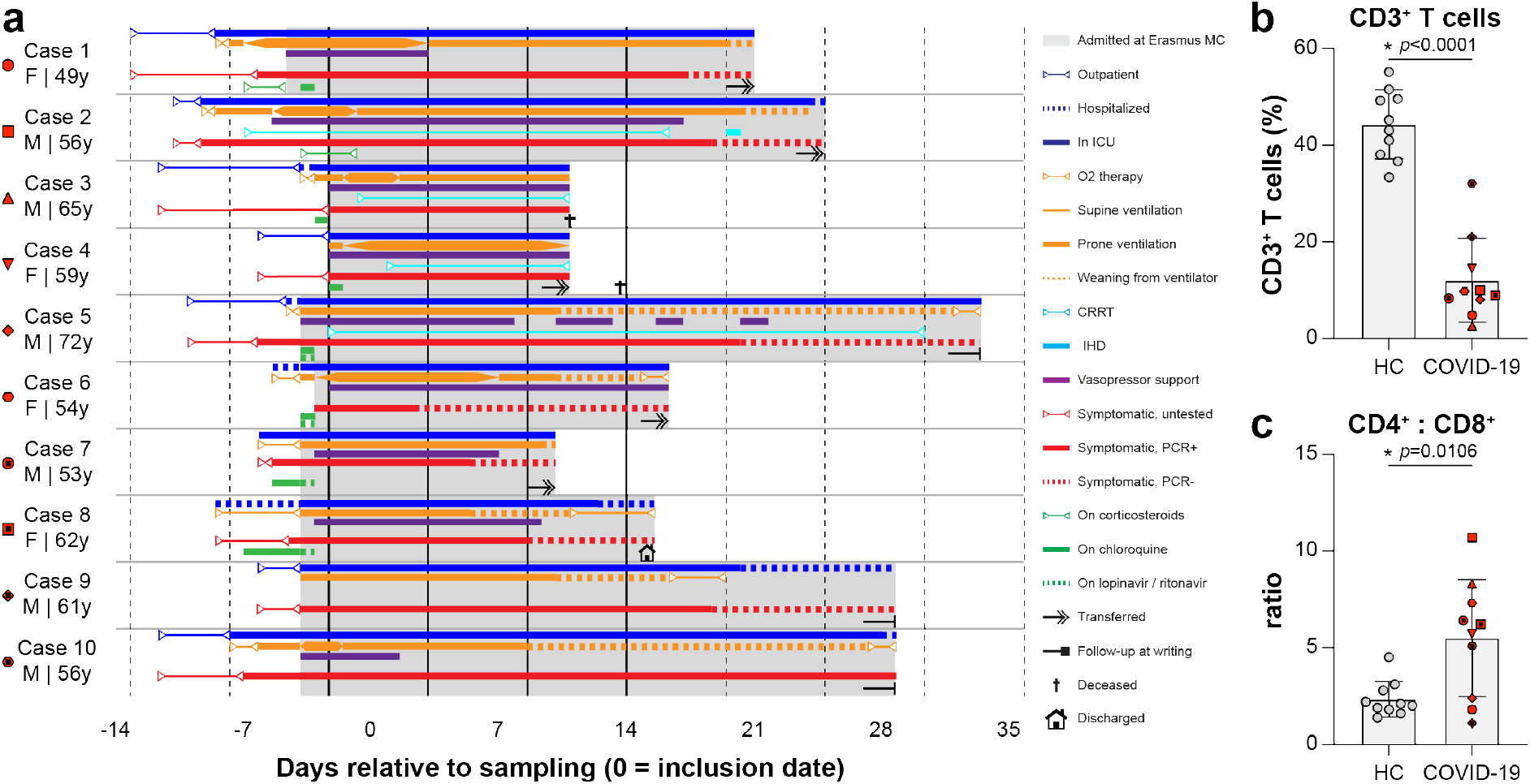
Clinical overview of moderate to severe COVID-19 ARDS patients. (a) Symptoms, hospitalization status, treatment and follow-up of n=10 COVID-19 ARDS patients included in this study. PBMC samples were obtained weekly after admission to the study. Symbols shown next to the cases match throughout all figures. (b) Flow cytometry performed on PBMC collected 14 days post inclusion showed that percentages CD3^+^ T-cells within the total LIVE gate were significantly lower in COVID-19 patients compared to healthy controls (HC), while (c) CD4:CD8 ratios were significantly higher. Panels b and c show individual values for n=10 patients versus n=10 HC, as well as the mean ± SD. Asterisk denotes a significant difference. HC = healthy control.

Phenotyping analysis of PBMC collected 14 days post inclusion via flow cytometry indicated that COVID-19 patients presented with low percentages of CD3^+^ T-cells in peripheral blood, corresponding to the previously reported lymphopenia (12.1 ± 8.7% in COVID-19 vs 44.3 ± 7.1% in HC, *p*<0.0001, Figure 1b) (Chen et al., 2020; Huang et al., 2020). CD4:CD8 ratios were increased in COVID-19 patients when compared to HC (5.5 ± 3.0 in COVID-19 vs 2.3 ± 0.9 in HC, *p*=0.0106, Figure 1c).

PBMC from COVID-19 ARDS patients were stimulated with three different peptide MPs: MP_S, MP_CD4_R and two MP_CD8 pools. MP_S contained 221 overlapping peptides (15-mers overlapping by 10 amino acids) covering the entire S glycoprotein and can stimulate both CD4^+^ and CD8^+^ T-cells. MP_CD4_R contained 246 HLA class II predicted epitopes covering all viral proteins except S, specifically designed to activate CD4^+^ T-cells. The two MP_CD8 pools combined contained 628 HLA class I predicted epitopes covering all SARS-CoV-2 proteins, specifically designed to activate CD8^+^ T-cells (Grifoni et al., 2020). Results obtained with MP_CD8_A and MP_CD8_B have been concatenated and shown as a single stimulation in Figure 2, results obtained with separate stimuli are shown in Supplementary Figure 2. In addition to stimulation of PBMC from COVID-19 ARDS patients, ten HC were included in this study as negative controls. PBMC from healthy controls were obtained before 2020 and could therefore not contain SARS-CoV-2-specific T-cells, however they potentially contain cross-reactive T-cells induced by circulating seasonal ‘common cold’ coronaviruses (Kissler et al., 2020).

**Figure 2.**
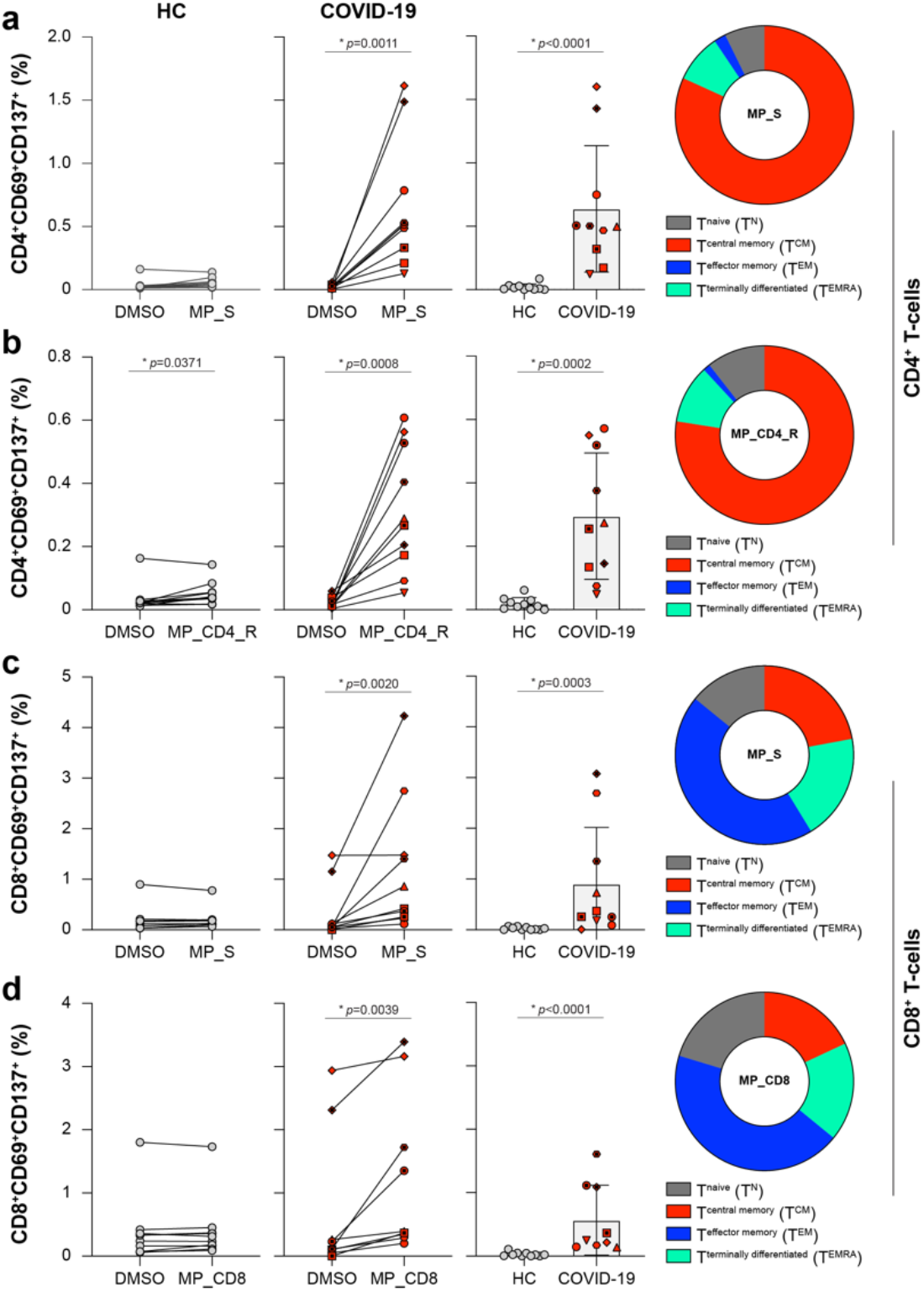
SARS-CoV-2-specific T-cell responses in COVID-19 ARDS patients. (a, b) Antigen-specific activation of CD4^+^ T-cells after stimulation for 20 hours with MP_S (a) and MP_CD4_R (b), measured via cell surface expression of CD69 and CD137 (gating in supplementary Figure 1). Two left panels show activation percentages (within CD3^+^CD4^+^ or CD3^+^CD8^+^ gate) obtained with the vehicle control (DMSO) and specific stimulation (MP) for HC and COVID-19 patients. The third panel shows the specific activation percentages corrected by subtracting the background present in the DMSO stimulation to allow comparison of both groups. The fourth panel shows the memory phenotype of the CD69^+^CD137^+^ responder cells in a donut diagram. (c, d) Similar layout to panels a and b. Antigen-specific activation of CD8^+^ T-cells after stimulation for 20 hours with MP_S (c) and MP_CD8 (d). Percentages from MP_CD8 stimulation are based on concatenated analysis of MP_CD8_A and MP_CD8_B, results obtained with separate stimulations are shown in supplementary Figure 2. Panels show individual values for n=10 patients versus n=10 HC, as well as the mean ± SD. Asterisk denotes a significant difference. HC = healthy control. Symbol shapes of COVID-19 patients are identical between panels, and refer back to figure 1.

Stimulation of PBMC collected 14 days post inclusion with the different peptide pools led to consistent detection of CD4^+^ and / or CD8^+^ SARS-CoV-2-specific T-cells in COVID-19 ARDS patients (Figure 2, Supplementary Figure 2 and Table 1). Specific activation of CD4^+^ and CD8^+^ T-cells was measured via cell surface expression of CD69 and CD137; phenotyping of memory subsets was based on surface expression of CD45RA and CCR7 (Supplementary Figure 1).

Stimulation of PBMC with MP_S and MP_CD4_R led to consistent activation of SARS-CoV-2-specific CD4^+^ T-cells (Figure 2a and b) in PBMC obtained from COVID-19 ARDS patients. Significant responses were detected when activation percentages after stimulation with MP_S and MP_CD4_R were compared with the vehicle control (DMSO). To allow comparison between HC and COVID-19 ARDS patients, we corrected the MP-specific activation percentages by subtracting the value obtained in the DMSO stimulation. Significant T-cell responses were observed in COVID-19 ARDS patients when compared with HC (0.64% in COVID-19 vs 0.02% in HC, *p*<0.0001 for MP_S and 0.29% in COVID-19 vs 0.02% in HC, p=0.0002 for MP_CD4_R, Figure 2a). The stimulation index (SI) was calculated by dividing the MP-specific responses by the DMSO responses (Supplementary Figure 2a), and donors with a SI > 3 were regarded responders (Table 1). According to this definition all COVID-19 ARDS patients responded to the MP_S and MP_CD4_R pools, whereas only 1/10 and 2/10 of the HC responded, respectively. Overall, the MP_S peptide pool induced stronger responses than the MP_CD4_R peptide pool, indicating that the S glycoprotein is a strong inducer of CD4^+^ T-cell responses. Phenotyping of CD4^+^CD69^+^CD137^+^ activated T-cells identified the majority of these SARS-CoV-2-specific T-cells as central memory T-cells, based on CD45RA and CCR7 expression (T^CM^). T^CM^ express homing receptors required for extravasation and migration to secondary lymphoid tissues, but also have high proliferative capacity with low dependence on co-stimulation (Mahnke et al., 2013; Sallusto et al., 1999).

**Table 1.** Antigen-specific responses shown as stimulation index (SI) for HC and COVID-19 patients. Antigen-specific activation of CD4^+^ and CD8^+^ T-cells in COVID-19 patients after stimulation for 20 hours with peptide MegaPools (MP) shown as stimulation index (SI). SI is derived by dividing the percentage obtained with specific stimulation (MP) by the percentage obtained with the vehicle control (DMSO), values for respective stimulations are shown in Figure 2. Donors with a SI > 3 are regarded responders to MP stimulation, total responders per stimulation are shown at the bottom of the table. Non-responders (SI < 3) are shown in white background, responders are shown in shades of blue (darker blue equals a higher SI). HC = healthy control.

| Activation (SI) |  |  |  |  |  |
| --- | --- | --- | --- | --- | --- |
|  |  | CD4+ T-cells |  | CD8+ T-cells |  |
| Paper ID |  | MP_S | MP_CD4_R | MP_S | MP_CD8 |
| HC | HC_1 | 2.0 | 1.7 | 1.0 | 1.0 |
|  | HC_2 | 0.8 | 0.9 | 0.9 | 1.0 |
|  | HC_3 | 0.9 | 0.9 | 1.6 | 1.0 |
|  | HC_4 | 1.7 | 1.9 | 1.1 | 1.1 |
|  | HC_5 | 1.4 | 1.3 | 3.0 | 2.6 |
|  | HC_6 | 8.3 | 3.6 | 3.1 | 1.8 |
|  | HC_7 | 1.4 | 1.4 | 0.9 | 1.1 |
|  | HC_8 | 1.1 | 2.6 | 1.2 | 1.1 |
|  | HC_9 | 2.4 | 3.8 | 1.8 | 1.3 |
|  | HC_10 | 1.7 | 1.4 | 1.0 | 0.9 |
| total |  | 1/10 | 2/10 | 1/10 | 0/10 |
| COVID-19 | Case 1 | 22.0 | 17.0 | 4.5 | 3.8 |
|  | Case 2 | 5.4 | 4.4 | 8.4 | ND |
|  | Case 3 | 33.4 | 18.9 | 6.7 | 1.5 |
|  | Case 4 | 29.8 | 12.6 | 4.5 | 3.4 |
|  | Case 5 | 123.3 | 43.0 | 1.0 | 1.1 |
|  | Case 6 | 28.2 | 5.4 | 48.0 | 2.5 |
|  | Case 7 | 57.4 | 58.8 | 3.2 | 5.8 |
|  | Case 8 | 28.6 | 23.0 | 1.7 | 1.2 |
|  | Case 9 | 25.2 | 3.5 | 3.7 | 1.5 |
|  | Case 10 | 18.9 | 14.4 | 25.8 | 15.8 |
| total |  | 10/10 | 10/10 | 8/10 | 4/9 |

SARS-CoV-2-specific CD8^+^ T-cells were activated by both the MP_S and MP_CD8 peptide pools when compared to vehicle control (Figure 2c and d). Furthermore, significant responses were detected when activation percentages after stimulation with MP_S and MP_CD8 were compared between HC controls and COVID-19 ARDS patients after DMSO correction (0.90% in COVID-19 vs 0.03% in HC, *p*=0.0003 for MP_S and 0.57% in COVID-19 vs 0.03% in HC, p<0.0001 for MP_CD8, Figure 2b). In addition to inducing specific CD4^+^ T-cells, the S glycoprotein also induced CD8^+^ T-cell responses. Calculation of the SI identified 8/10 and 4/9 of the COVID-19 ARDS patients as responders to MP_S and MP_CD8, respectively, whereas 1/10 of the HC responded to the MP_S stimulation (Supplementary Figure 2, Table 1). Phenotyping of CD8^+^CD69^+^CD137^+^ activated T-cells showed that these had a mixed phenotype. The majority of virus-specific CD8^+^ T-cells was identified as CCR7^-^ effector memory (T^EM^) or terminally differentiated effector (T^EMRA^) (Mahnke et al., 2013). Both these CD8^+^ effector subsets are potent producers of IFNγ, contain preformed perforin granules for immediate antigen-specific cytotoxicity and home efficiently to peripheral lymphoid tissues (Sallusto et al., 1999).

As production of pro-inflammatory cytokines can be predictive of clinical outcome for other viral diseases (Wang et al., 2014), we measured antigen-specific production of 13 cytokines in cell culture supernatants from PBMC after stimulation. PBMC were stimulated with the respective peptide pools, cytokine production after MP_S stimulation is shown in Figure 3 and Supplementary Figure 3 as representative data (data for other stimulations not shown). When compared to the vehicle control stimulation, PBMC obtained from COVID-19 ARDS patients specifically produced IFNγ, TNFα, IL-2, IL-5, IL-13, IL-10, IL-9, IL-17a, IL-17f and IL-22 after MP_S stimulation (Figure 3, Supplementary Figure 3).

When comparing COVID-19 ARDS patients with HC, stimulation of PBMC by the overlapping S peptide pool led to a strong significant production of the Th1 or effector cytokines IFNγ, TNFα and IL-2 in COVID-19 ARDS patients. More characteristic Th2 cytokines (IL-5, IL-13, IL-9 and IL-10) were also consistently detected, albeit at low levels. IL-4 and IL-21 could not be detected at all. IL-6 levels were not different between COVID-19 patients and HC, however results were difficult to interpret because mock stimulation already resulted in high IL-6 expression. Antigen-specific production of cytokines related to a Th17 response was mixed; PBMC from COVID-19 ARDS patients produced significantly more IL-17a and IL-22 than HC; this was not observed for IL-17f.

**Figure 3.**
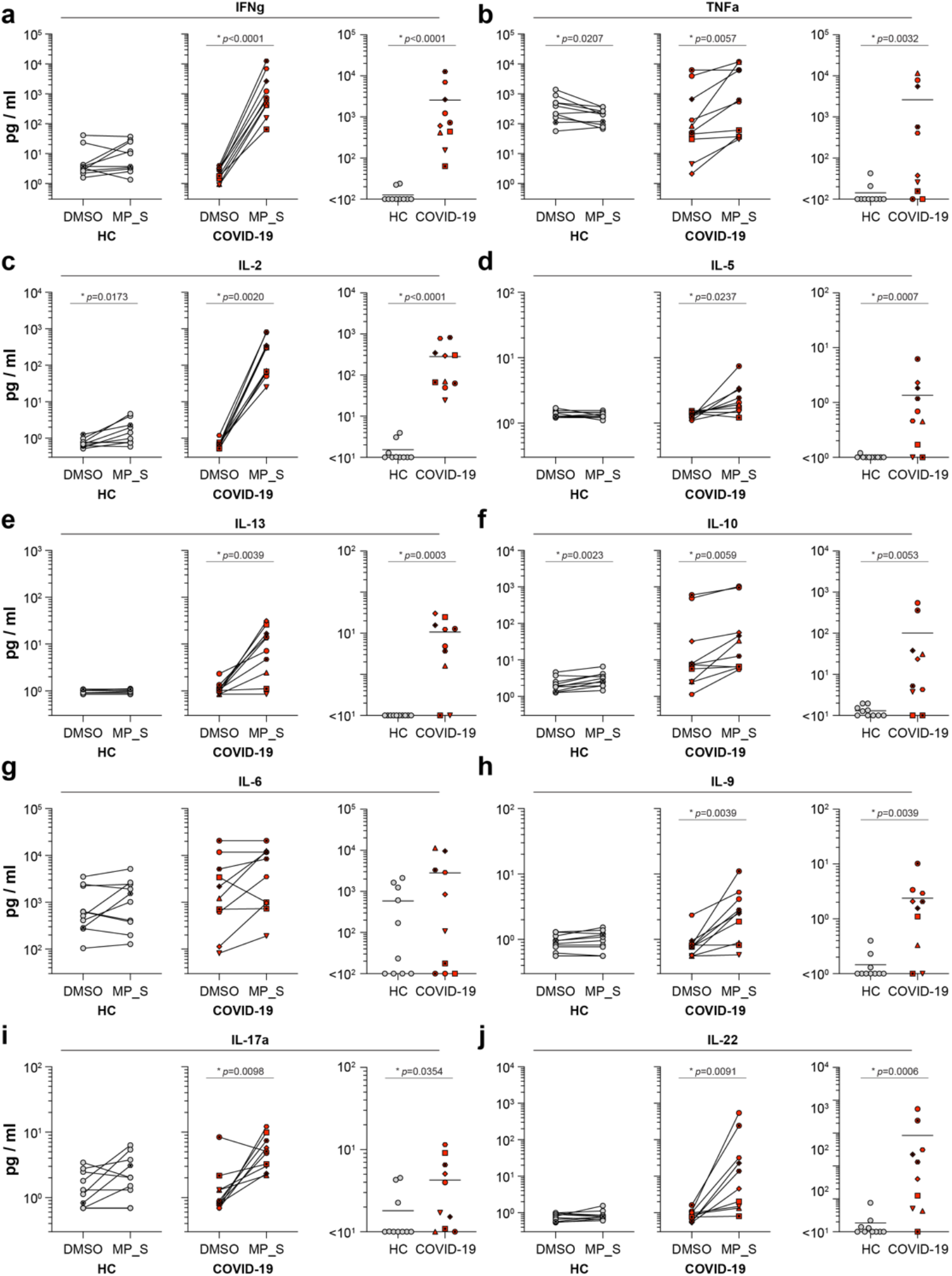
SARS-CoV-2-specific cytokine production in COVID-19 ARDS patients. (a-j) Antigen-specific production of cytokines measured in cell culture supernatants from PBMC stimulated (20 hours) with MP_S. Two left panels show activation percentages obtained with the vehicle control (DMSO) and specific stimulation (MP) for HC and COVID-19 patients. The third panel shows the quantity corrected by subtracting the background present in the DMSO stimulation to allow comparison of both groups. Panels show individual values for n=10 patients versus n=10 HC, as well as the geometric mean. Asterisk denotes a significant difference. HC = healthy control. Additional cytokines (IL-4, IL17f and IL-21) are shown in supplementary Figure 3.

In general, stimulation of PBMC from COVID-19 ARDS patients with MP led to a dominant production Th1 or effector cytokines (IFNγ, TNFα, IL-2), but Th2 (IL-5, IL-13, IL-9, IL-10) and Th17 cytokines could also be detected (IL-17a, IL-17f and IL-22). Although not enough COVID-19 ARDS patients were included in this study to correlate specific T-cell responses to clinical outcome, we did observe differences in cytokine production profiles on a case-per-case basis (Supplementary Figure 3d). Plotting the respective cytokine quantities as a percentage of total cytokine production showed that either IL-6 (case 3, 5 and 9), TNFα (case 1, 3 and 9), IL-2 (case 8) or IFNγ (case 2, 4, 6, 7 and 10) dominated the response.

Finally, we studied the kinetics of development of virus-specific humoral and cellular immune response in eight COVID-19 ARDS patients included in this study (case 1 – 8). Real time RT-PCR detection of SARS-CoV-2 genomes in respiratory tract samples showed a decreasing trend over time (Figure 4a, ANOVA repeated measures *p*<0.001), whereas virus-specific serum IgG antibody levels, measured by RBD ELISA, showed a significant increase (Figure 4b, ANOVA repeated measures, *p*<0.001). Concomitantly, SARS-CoV-2-specific T-cells were detected in all patients at multiple time-points and frequencies of virus-specific responder cells increased significantly over time (Figure 4c, ANOVA repeated measures, *p*<0.001).

**Figure 4.**
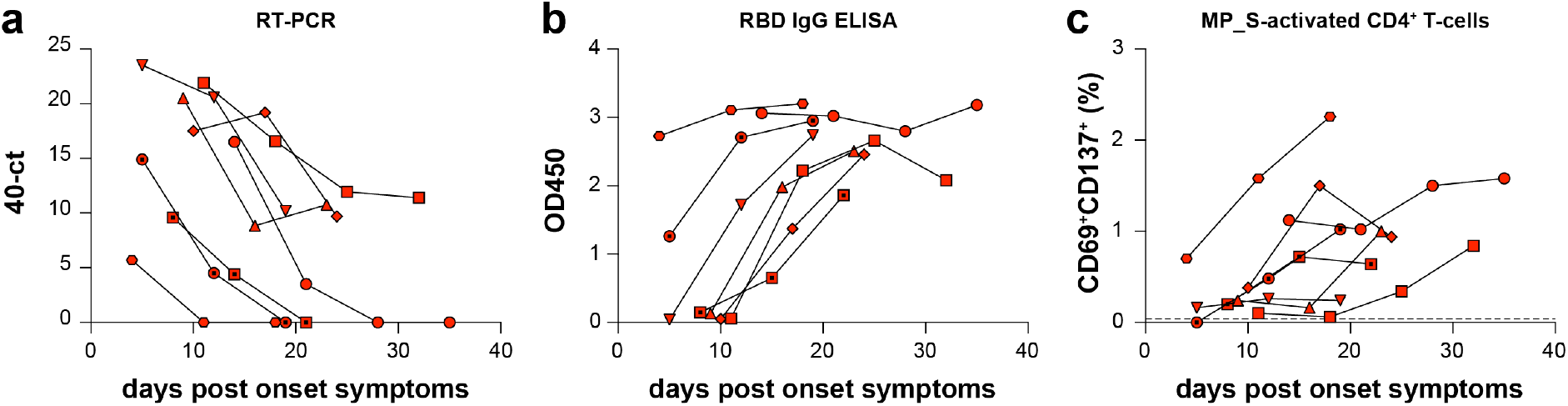
SARS-CoV-2 replication and humoral and cellular immune response kinetics in COVID-19 ARDS patients. (a, b, c) Sequential measurements of SARS-CoV-2 genomes detected in upper respiratory tract samples by real-time RT-PCR (40-ct, a), SARS-CoV-2-specific serum RBD IgG antibody levels detected by ELISA (OD450, b) and percentage SARS-CoV-2-specific CD4^+^ T-cells after MP_S stimulation of PBMC (c), plotted against days post onset of symptoms. Genome levels showed a significant decrease over time, antibody levels and specific T-cell frequencies significantly increased (*p*<0.001 for all three panels, ANOVA repeated measures). Panels show values for n=8 COVID-19 ARDS patients (case 1 – 8).

Collectively, these data provide information on the phenotype, breadth and kinetics of virus-specific cellular immune responses in COVID-19 ARDS patients. We provide evidence that SARS-CoV-2-specific CD4^+^ and CD8^+^ T-cells appear in blood of ARDS patients in the first two weeks post onset of symptoms, and their frequency increases over time. SARS-CoV-2-specific CD4^+^ T-cells in blood typically had a central memory phenotype, whereas CD8^+^ T-cells had a more effector phenotype. Consistent production in response to viral antigen of IFNγ, TNFα, IL-2, IL-5, IL-13, IL-9, IL-10, IL-17a, IL-17f and IL-22 was observed, with a dominant production of the effector and Th1 cytokines. Due to limitations in the number of PBMC that could be obtained from severe COVID-19 ARDS patients in an ICU setting, we could not resolve which cells were responsible for production of which cytokine by intracellular cytokine staining.

Elevated levels of IL-6 in patient plasma have been correlated to respiratory failure in COVID-19 patients (Herold et al., 2020). Although we could not detect increased specific production of IL-6 in PBMC stimulated with peptide pools due high background production in controls (potentially due to *in vivo* activation), we detected a dominant IL-6 and TNFα response in cell culture supernatants from the patient deceased due to respiratory failure (case 3). To determine the role of T-cells in COVID-19, it is crucial that the cell types responsible for the production of IL-6 and the concomitant ‘cytokine storm’ are identified in large comparative cohort studies.

We included PBMC obtained from ten buffycoats obtained before the SARS-CoV-2 pandemic as negative HC. In some instances, reactive T-cells were detected in HC after MP stimulation, both on basis of T-cell activation and cytokine production. Since PBMC from these HC could not contain SARS-CoV-2-specific T-cells, we hypothesize that these responses were cross-reactive and had been induced by circulating seasonal ‘common cold’ coronaviruses. If we consider samples with a SI > 3 as responders, we identified 2 out of 10 HC (20%) to have these cross-reactive T-cells. This is in good accordance with a recent report on the presence of virus-specific CD4^+^ T-cells in COVID-19 patients, which reported the presence of SARS-CoV-2-reactive T-cells in 34% of the HC (Braun et al., 2020). The role of pre-existing SARS-CoV-2-reactive T-cells as a correlate of protection or pathology is unclear, and needs to be addressed in prospective studies.

Novel SARS-CoV-2 vaccines are currently in development and mainly focus on the surface glycoprotein S as an antigen for efficient induction of virus-specific neutralizing antibodies. We now show that S can also be a potent immunogen for inducing virus-specific CD4^+^ and CD8^+^ T-cells. Whether presence and certain phenotypes of T-cells are correlated to a ‘good’ or ‘bad’ prognosis remains to be determined. Collectively, these novel data are important for vaccine design and will facilitate the evaluation of future vaccine immunogenicity.

## Data Availability

All data generated or analysed during this study are included in this manuscript (and its supplementary information files).

## Author contributions

DW, AG, RLdS, AS and RDdV conceived and planned the experiments. DW, KSS, MPR, NMAO, RM and ECMvG contributed to sample preparation. DW, KSS, MPR, AG, NMAO, HE, JPCvsA, RM and RDdV carried out the experiments. DW, KSS, AG, MPGK, BLH, RLdS, AS and RDdV contributed to the interpretation of the results. RDdV took the lead in writing the manuscript, DW, KSS and RLdS contributed significantly. All authors provided critical feedback and helped shape the research, analysis and manuscript.

## Acknowledgements

We thank all health care workers and laboratory personnel who contributed to treatment and diagnosis of these and other COVID-19 patients. Specifically, we thank Jeroen van Kampen, Corine Geurts van Kessel, Annemiek van der Eijk and Marshall Lammers for their contributions to these studies. This work has received funding from the European Union’s Horizon 2020 research and innovation programme under grant agreements No. 874735 (VEO) (MPGK, EvG and MPR).

## Supplementary Figure

**Supplementary Figure 1.**
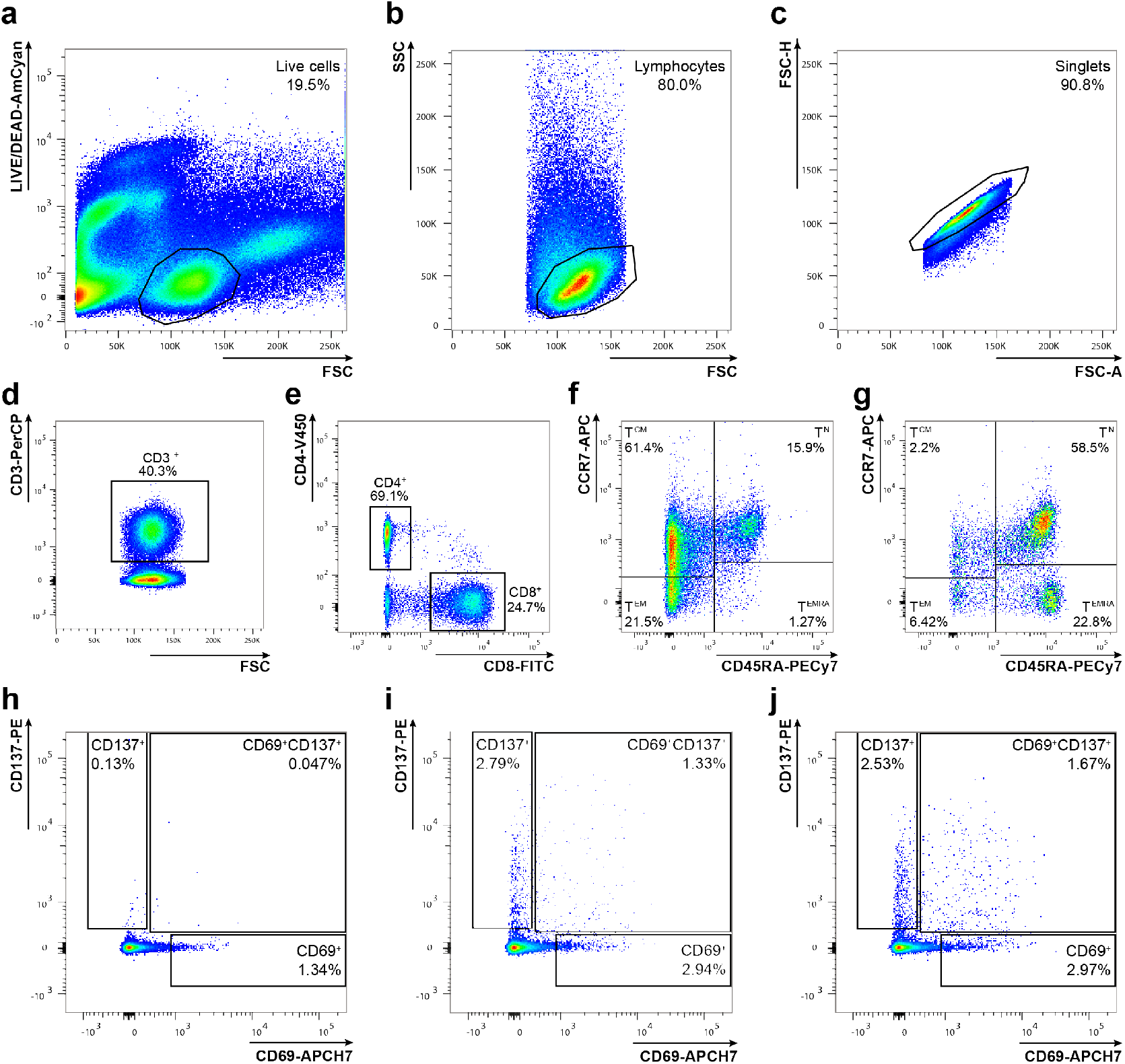
Flow cytometry gating strategy. (a-j) Gating strategy for detection of antigen-specific T-cells after stimulation of PBMC. (a) Live cells were selected, (b) followed by the selection of lymphocytes and (c) singlets. (d) CD3^+^ T-cells were gated and (e) divided into CD4^+^ and CD8^+^ T-cells. Both subsets were phenotyped as naïve (T^N^), central memory (T^CM^), effector memory (T^EM^) or terminally differentiated effectors (T^EMRA^) on basis of expression of CD45RA and CCR7 (f for CD4^+^ T-cells, g for CD8^+^ T-cells). (h-j) Within the different subsets, activated cells were identified via surface upregulation of activation induced markers CD69 and CD137. Percentages of CD69^+^CD137^+^ double positive cells, reflecting activated cells, were used for further analysis. Representative examples of DMSO (vehicle of the MP) (background control, h), CMV peptides (positive control, i) and a MP_S stimulation of PBMC from a COVID-19 patient.

**Supplementary Figure 2.**
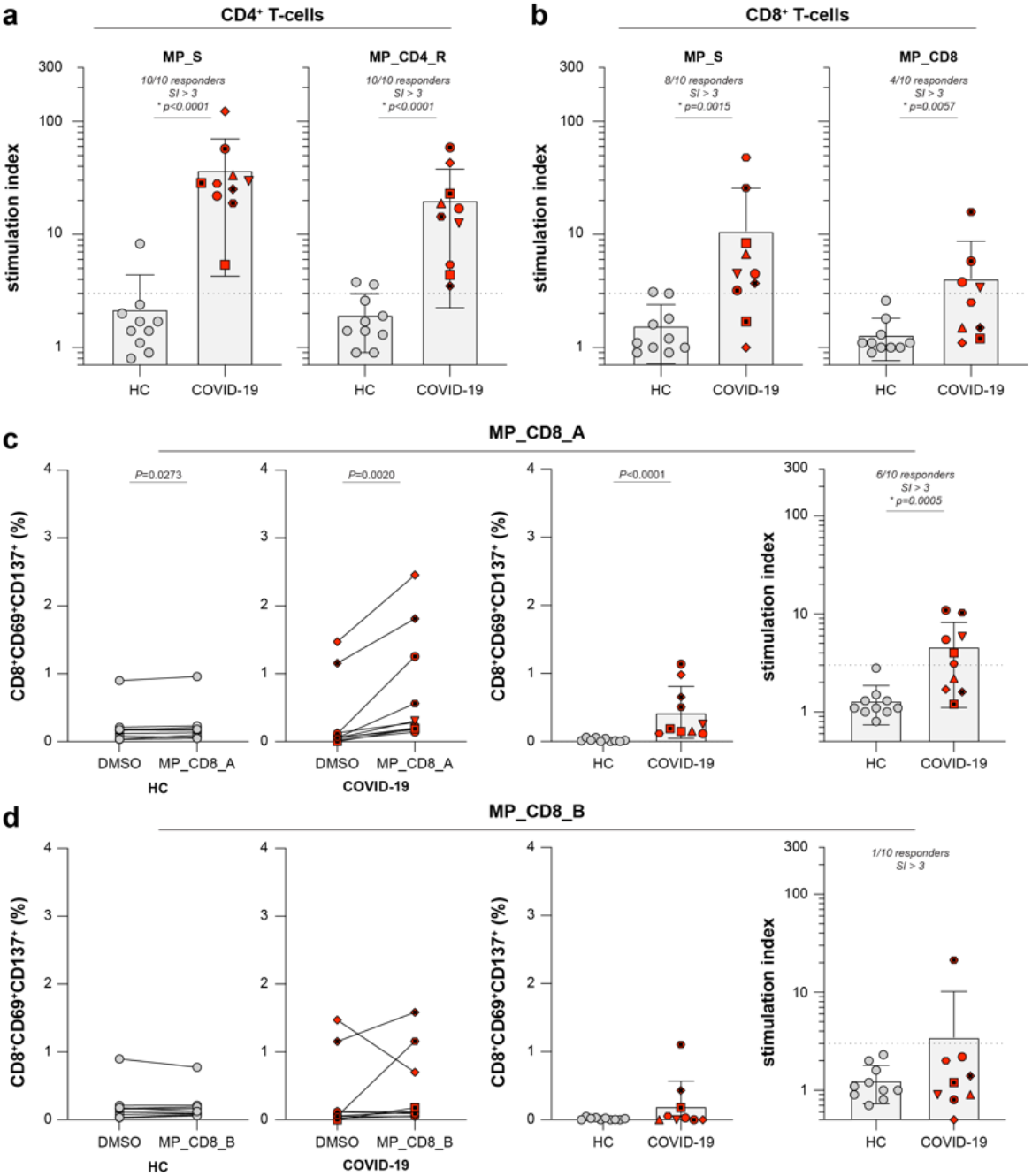
SARS-CoV-2-specific T-cell responses in COVID-19 ARDS patients. (a, b) Antigen-specific activation of CD4^+^ and CD8^+^ T-cells in COVID-19 patients after stimulation for 20 hours with peptide MegaPools (MP) shown as stimulation index (SI). Stimulation index is derived by dividing the percentage obtained with specific stimulation (MP) by the percentage obtained with the vehicle control (DMSO), values for respective stimulations are shown in Figure 2. Donors with a SI > 3 (dotted line) are regarded responders to MP stimulation. (c, d) Antigen-specific activation of CD8^+^ T-cells after stimulation for 20 hours with MP_CD8_A (c) and MP_CD8_B (d). The two left panels show activation percentages obtained with the vehicle control (DMSO) and specific stimulation (MP) for HC and COVID-19 patients. The third panel shows the specific activation percentages corrected by subtracting the background present in the DMSO stimulation to allow comparison of both groups. The fourth panel shows the stimulation index derived by dividing the percentage obtained with specific stimulation (MP) by the percentage obtained with the vehicle control (DMSO). Donors with a SI > 3 (dotted line) are regarded responders to MP stimulation. Panels show individual values for n=10 patients versus n=10 HC, as well as the mean ± SD. Asterisk denotes a significant difference. HC = healthy control.

**Supplementary Figure 3.**
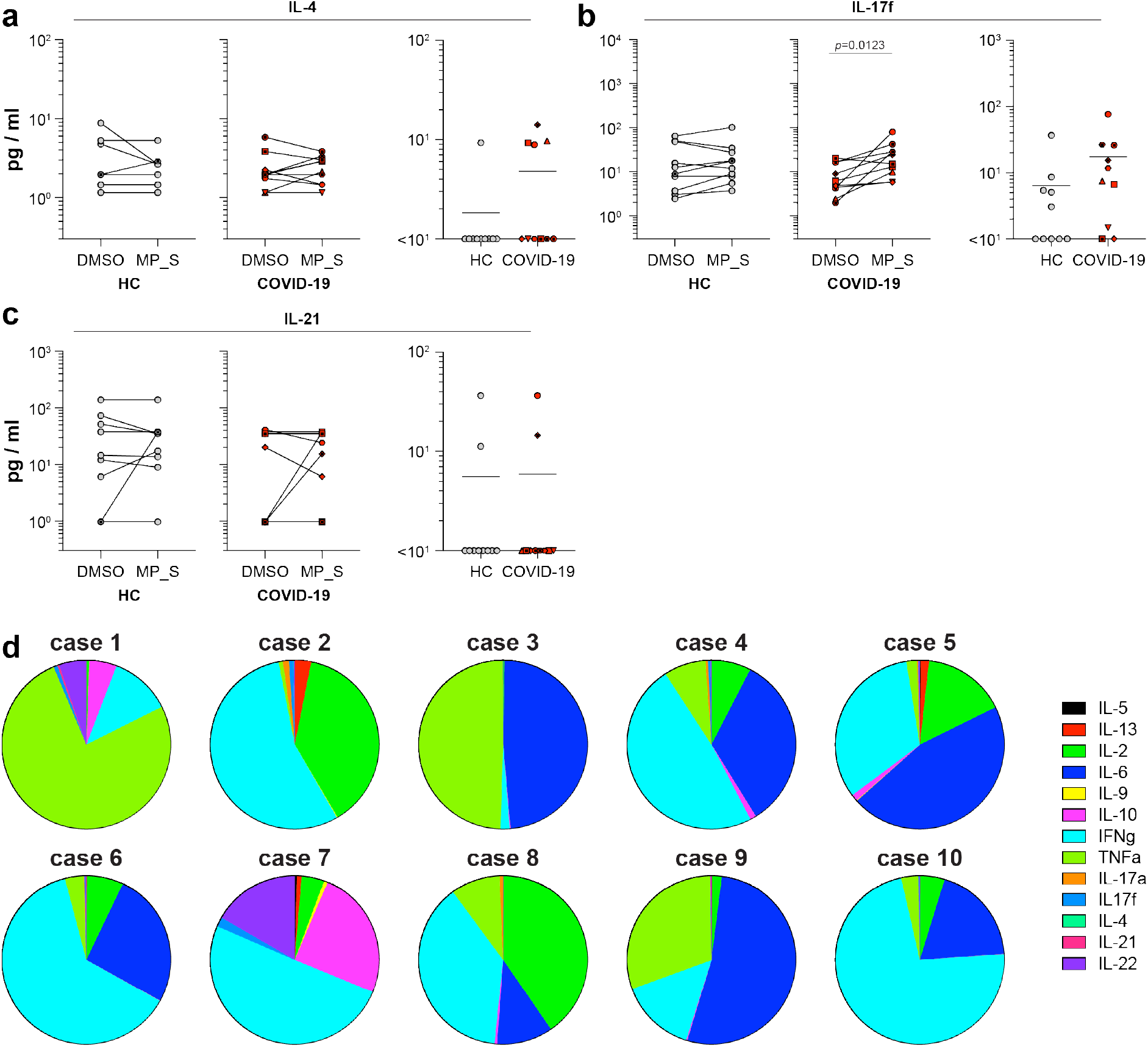
SARS-CoV-2-specific cytokine production in COVID-19 ARDS patients. (a-c) Antigen-specific production of cytokines measured in cell culture supernatants from PBMC stimulated (20 hours) with MP_S. Two left panels show activation percentages obtained with the vehicle control (DMSO) and specific stimulation (MP) for HC and COVID-19 patients. Third panel shows the quantity corrected by subtracting the background present in the DMSO stimulation to allow comparison of both groups. Panels show individual values for n=10 patients versus n=10 HC, as well as the geometric mean. Asterisk denotes a significant difference. HC = healthy control. (d) Antigen-specific production of cytokines per COVID-19 case. Circle diagrams represent the total amount of cytokines produced by the respective donor (corrected for DMSO background), quantities of different cytokines are shown as a percentage of whole. Clinical data for cases is described in Figure 1.

## Methods

### Ethical statement

Patients admitted into the intensive care unit (ICU) with Acute Respiratory Distress Syndrome (ARDS) resulting from SARS-CoV-2 infection at Erasmus MC, Rotterdam, the Netherlands were included in a biorepository study aimed at ARDS and sepsis in the ICU. The first EDTA blood samples for PBMC isolation were obtained no more than 2 days after admission into the Erasmus MC ICU until 21 days post inclusion for as long as the patient was in the ICU. Patient care and research were conducted in compliance within guidelines of the Erasmus MC and the Declaration of Helsinki. Due to the clinical state of most ARDS patients (*i.e*. intubated, comatose), deferred proxy consent was obtained instead of direct written informed consent from the patients themselves. Retrospective written informed consent was obtained from patients after recovery. The study protocol was approved by the medical ethical committee of Erasmus MC, Rotterdam, the Netherlands (MEC-2017-417 and MEC-2020-0222). For experiments involving healthy control human buffy coats, written informed consent for research use was obtained by the Sanquin Blood Bank (Rotterdam, the Netherlands).

### Diagnosis

Real-time RT-PCR on the E-gene was performed as described previously (Corman et al., 2020) on RNA isolated from sputa, nasopharyngeal or oropharyngeal swabs by MagnaPure (Roche diagnostics, The Netherlands) using the total nucleic acid (TNA) isolation kit.

### PBMC isolation

PBMC were isolated from EDTA blood samples. Tubes were centrifuged at 200g for 15 min to separate cellular parts. Plasma-containing fraction was collected, centrifuged at 1200g for 15 minutes, plasma was aliquoted and stored at −20°C. The cellular fraction was reconstituted with phosphate-buffered saline (PBS) and subjected to Ficoll density gradient centrifugation (500g, 30min). PBMC were washed and frozen in 90% fetal bovine serum (FBS) and 10% dimethyl sulfoxide (DMSO, Sigma Life science) at −135°C. Upon use, PBMC were thawed in IMDM (Lonza, Belgium) supplemented with 10% FBS, 100 IU of penicillin/ml, 100 μg of streptomycin/ml (Lonza, Belgium) and 2 mM L-glutamine (Lonza, Belgium) (I10F medium). PBMC were treated with 50 U/ml benzonase (Merck) for 30min at 37°C prior to use in stimulation assays.

### Epitope MegaPool (MP) design and preparation

SARS-CoV-2 virus-specific CD4 and CD8 peptides were synthesized as crude material (A&A, San Diego, CA), resuspended in DMSO, pooled and sequentially lyophilized as previously reported (Carrasco Pro et al., 2015). SARS-CoV-2 epitopes were predicted using the protein sequences derived from the SARS-CoV-2 reference (GenBank: MN908947) and IEDB analysis-resource as previously described (Dhanda et al., 2019; Grifoni et al., 2020). Specifically, CD4 SARS-CoV-2 epitope prediction was carried out using a previously described approach in Tepitool resource in IEDB (Paul et al., 2015; Paul et al., 2016) similarly to what was previously described (Grifoni et al., 2020), but removing the resulting Spike glycoprotein epitopes from this prediction (CD4-R(remainder) MP, n=246). To investigate in depth Spike-specific CD4^+^ T-cells, overlapping 15-mer by 10 amino acids have been synthesized and pooled separately (CD-4 S(spike) MP, n=221). CD8 SARS-CoV-2 epitope prediction was performed as previously reported, using the NetMHCpan4.0 algorithm for the top 12 more frequent HLA alleles in the population (HLA-A*01:01, HLA-A*02:01, HLA-A*03:01, HLA-A*11:01, HLA-A*23:01, HLA-A*24:02, HLA-B*07:02, HLA-B*08:01, HLA-B*35:01, HLA-B*40:01, HLA-B*44:02, HLA-B*44:03) and selecting the top 1 percentile predicted epitope per HLA allele (Grifoni et al., 2020). The 628 predicted CD8 epitopes were split in two CD8 MPs containing 314 peptides each.

### SARS-CoV-2 RBD ELISA

Serum or plasma samples were analyzed for the presence of SARS-CoV-2 specific antibody responses using a validated in-house SARS-CoV-2 receptor binding domain (RBD) IgG ELISA as previously described(Okba et al., 2020). Briefly, ELISA plates were coated with recombinant SARS-CoV-2 RBD protein. Following blocking, samples were added and incubated for 1 hour, after which the plates were washed and a secondary HRP-labelled rabbit anti-human IgG (DAKO) was added. Following a 1 hour incubation, the plates were washed, the signal was developed using TMB, and the OD_450_ was measured for each well. All samples reported here were interrogated for the presence of antibodies on the same plate.

### *Ex vivo* stimulations

PBMC were plated in 96-wells U bottom plates at 1 × 10^6^ PBMC per well in RPMI1640 (Lonza, Belgium) supplemented with 10% human serum, 100 IU of penicillin/ml, 100 μg of streptomycin/ml (Lonza, Belgium) and 2 mM L-glutamine (Lonza, Belgium) (R10H medium) and subsequently stimulated with the described CD4 and CD8 SARS-CoV-2 MPs at 1μg/ml. A stimulation with an equimolar amount of DMSO was performed as negative control, phytohemagglutinin (PHA, Roche, 1μg/ml) and stimulation with a combined CD4 and CD8 cytomegalovirus MP (CMV, 1μg/ml) were included as positive controls. Twenty hours after stimulation cells were stained for detection of activation induced markers and subjected to flow cytometry. Supernatants were harvested for multiplex detection of cytokines.

### Flow cytometry

Activation-induced markers were quantified via flow cytometry (FACSLyric, BD Biosciences). A surface staining on PBMC was performed with anti-CD3^PerCP^ (BD, clone SK7), anti-CD4^V450^ (BD, clone L200), anti-CD8^FITC^ (DAKO, clone DK25), anti-CD45RA^PE-Cy7^ (BD, clone L48), anti-CCR7^APC^ (R&D Systems, clone 150503), anti-CD69^APC-H7^ (BD, clone FN50) and anti-CD137^PE^ (Miltenyi, clone 4B4-1). T-cell subsets were identified via the following gating strategy: LIVE CD3^+^ were selected and divided in CD3^+^CD4^+^ and CD3^+^CD8^+^. Within the CD4 and CD8 subsets, memory subsets were gated as CD45RA^+^CCR7^+^ (naive, T^n^), CD45RA^-^, CCR7^+^ (central memory, T^CM^), CD45RA^-^CCR7^-^ (effector memory, T^EM^) or CD45RA^+^CCR7^-^ (terminally differentiated effectors, T^EMRA^). T-cells specifically activated by SARS-CoV-2 were identified by upregulation of CD69 and CD137. An average of 500,000 cells was always acquired, the gating strategy is schematically represented in (Supplementary Figure 1A-J). In analysis, PBMC stimulated with MP_CD8_A and MP_CD8_B were concatenated and analyzed as a single file for SARS-CoV-2-specific responses to MP_CD8.

### Multiplex detection of cytokines

Cytokines in cell culture supernatants from *ex vivo* stimulations were quantified using a human Th cytokine panel (13-plex) kit (LEGENDplex, Biolegend). Briefly, cell culture supernatants were mixed with beads coated with capture antibodies specific for IL-5, IL-13, IL-2, IL-6, IL-9, IL-10, IFNγ, TNFα, IL-17a, IL-17F, IL-4, IL-21 and IL-22 and incubated for 2 hours. Beads were washed and incubated with biotin-labelled detection antibodies for 1 hour, followed by a final incubation with streptavidin^PE^. Beads were analyzed by flow cytometry. Analysis was performed using the LEGENDplex analysis software v8.0, which distinguishes between the 13 different analytes on basis of bead size and internal dye. Quantity of each respective cytokine is calculated on basis of intensity of the streptavidin^PE^ signal and a freshly prepared standard curve.

### Statistical analysis

For comparison of CD3^+^ T-cell percentages, CD4:CD8 ratios, CD69^+^CD137^+^ stimulated T-cells and cytokine levels between HC and COVID-19 patients all log transformed data was tested for normal distribution. If distributed normally, groups were compared via an unpaired T test. If not distributed normally, groups were compared via a Mann-Whitney test. Comparisons between different stimulations (DMSO versus MP) were performed by paired T-test (normal distribution) or Wilcoxon rank test (no normal distribution). Two-tailed *p* values are reported throughout the manuscript. One way ANOVA repeated measures was used to test for increasing or decreasing trends over sequential time points (0, 7 and 14 days post inclusion).

